# Quantifying Effects of Lifestyle Changes on Progression to Advanced Age-Related Macular Degeneration in High Genetic Risk Individuals

**DOI:** 10.1101/2025.06.18.25329367

**Authors:** Johanna M. Seddon, Dikha De, Bernard Rosner

## Abstract

**Purpose:** We examined the extent to which adopting healthy lifestyle behaviors could offset high genetic risk for progression to advanced age-related macular degeneration (AMD), to address concerns of family members of affected patients.

**Design:** Prospective longitudinal study

**Participants:** Eyes with early or intermediate AMD at baseline were defined based on the Age-Related Eye Disease Study severity scale. High genetic risk was defined as the third tertile of a genetic risk score for progression, adjusted for age, race and sex.

**Methods:** Information on lifestyle behaviors was obtained from baseline risk and food frequency questionnaires. Risk-inducing and health-promoting lifestyle profiles were defined based on dichotomous categorizations of smoking, body-mass index (BMI), and dietary caloric intake, green leafy vegetables and fish, in never and ever smokers. Cox proportional hazard ratios (HRs), relative risks (RRs) and population attributable risks (PARs) were calculated, adjusting for inter-eye correlation, demographic factors, macular status and family history.

**Main Outcome Measures:** Progression to advanced AMD (AAMD) and subtypes geographic atrophy (GA) and neovascular (NV), confirmed at 2 consecutive visits over 5 years of follow-up.

**Results:** Among 898 high genetic risk eyes, 207 eyes progressed to AAMD (23%). Among never smokers, a high risk-inducing lifestyle profile conferred a 3-fold increased incidence of AAMD, compared to an ideal health-promoting lifestyle profile [HR = 3.3 (CI 1.8, 6.4), P <0.001]. In ever smokers, a risk-inducing profile was independently associated with a 5-fold increased incidence of AAMD [HR = 5.3 (CI 2.3,11.9), P <0.001]. Stronger effects of these lifestyle behaviors were seen for GA compared to NV. Estimated PARs suggested adopting an ideal health-promoting profile could prevent 56% of incident AAMD in never smokers and 60% in ever smokers.

**Conclusion:** Unhealthy behaviors increased incidence of AAMD by 3 to 5-fold among a highly genetically susceptible population, and 56-60% of AAMD incidence was attributed to the modifiable factors of smoking, high BMI, high caloric intake and low intake of foods rich in lutein-zeaxanthin and omega-3 fatty acids. These results underscore the importance of lifestyle interventions even in high genetic risk populations, such as relatives of affected patients, to reduce progression from early and intermediate AMD to advanced vision-threatening stages.

## INTRODUCTION

Age-related macular degeneration (AMD) is a multifactorial, neuro-degenerative disease which can cause irreversible vision loss in the elderly. Genetic factors contribute significantly to AMD, with heritability estimates ranging from 46%-67% in early or intermediate disease and higher heritability of 71% in advanced stages. ^1^ In addition to the association between genetics and disease prevalence, we have known since 2007 that genetic risk is a strong predictor of longitudinal progression from early and intermediate AMD to advanced stages. ^2^ Modifiable lifestyle factors including nutrition and smoking also play an important role, therefore both nature and nurture contribute to this disease. ^3–8^ Various diet-gene-lifestyle interactions have been reported to be associated with advanced AMD (AAMD) including associations between the complement factor H (*CFH*) Y402H genotype and body-mass index (BMI) ^9^ as well as the Mediterranean diet. ^10^ Effects of interactions between the age-related maculopathy susceptibility 2/high-temperature requirement A serine peptidase 1 (*ARMS2/HTRA1*) gene and dietary omega-3 fatty acid intake ^11^ and between complement component 3 (*C3*) R102G and folate intake ^12^ have also been reported for progression to geographic atrophy (GA).

Over the past decades, we have developed several predictive models to assess their impact on the likelihood of progression to advanced disease. ^8,13–16^ A recent model incorporating baseline demographic and behavioral factors, macular phenotype, family history of AMD and genetic variants was effective in discriminating eyes which progressed from no, early or intermediate stages to AAMD compared to non-progressing eyes over a 5 and 12 year follow-up period.^15^ Building on this model, we added dietary intake of green leafy vegetables and fatty fish to determine the effect of the composite model on transitions among the different AMD stages over 5 years. We found that increased consumption of foods rich in lutein-zeaxanthin and omega-3 fatty acids during the initial non-advanced AMD stages could reduce subsequent progression to higher non-advanced and advanced AMD severity stages.^16^ In all of these prediction models, higher risk scores were associated with higher rates of progression. Other reports have shown joint effects of lifestyles and genetic risk score on prevalence of AMD ^17,18^, and various forms of AMD progression and genetic risk defined by 2 genes.^19,20^ However to date, analyses targeted to the highest genetic risk population defined by a polygenic risk score, to determine the effect of a combination of diet and other lifestyle behaviors on progression to AAMD, GA and neovascular AMD (NV) have not been reported.

It would be helpful to expand our knowledge about whether adopting healthy lifestyle behaviors could offset very high genetic risk for progression to advanced stages of disease, since this information could be relevant in clinical practice. When patients with AMD ask about why they developed the disease, clinical providers can now explain the contributing factors and emphasize the strong genetic association, the link to family history, and the influence of modifiable lifestyle factors such as smoking, BMI, and diet.^8^ Accompanying family members often express concern about their own risk of developing AMD and may ask that since they are likely to have a high genetic predisposition, which is a decisive factor in AMD onset and progression, if they are destined to develop the disease.

In this study, we aimed to address this question and evaluated whether a high polygenic risk could be modified by adopting a healthy lifestyle. We examined the impact of a healthy lifestyle on the progression from early or intermediate AMD to AAMD in a cohort with high genetic susceptibility. Additionally, we quantified the risk associated with unhealthy behaviors and estimated the proportion of AAMD cases that could be prevented through lifestyle changes in individuals with a high genetic predisposition.

## METHODS

### Study population and definition of high genetic risk group

The Age-Related Eye Disease Study (AREDS) was a prospective multisite randomized clinical trial to evaluate the effect of antioxidants and mineral supplements on the risk of AMD and cataracts. ^21^ Participants were randomized to one of three treatment groups (antioxidants only, zinc only, or zinc + antioxidants combined) or a placebo group. Participants were aged 55 to 80 years at baseline and were required to have at least one eye with a visual acuity of no worse than 20/32. Clinic visits occurred every 6 months. Trained fundus graders, masked to clinical and phenotypic information from previous years, ascertained signs of AMD from annual stereoscopic color images by using a standardized and validated protocol at a single reading center. In this post-hoc analyses of AREDS data we used the severity scale ^22^ to classify eyes into severity group 1 (scale 1, no AMD), group 2 (scales 2-4, early AMD), group 3 (scales 5-8, intermediate AMD), and group 4 [scales 9-12, advanced AMD including both central and non-central forms of geographic atrophy (GA), and neovascular disease (NV)]. Eyes with AMD severity group 1 at baseline were excluded from the analyses.

Deidentified data were downloaded from the National Institutes of Health Database of Genotype and Phenotypes through accession number phs000001.v3.p1. Research adhered to the tenets of the Declaration of Helsinki performed under approved institutional review board protocols of the participating sites. This trial was registered at clinicaltrials.gov as NCT00594672.

The selection of subjects/eyes for this analysis is depicted in **Figure 1**. Eyes with baseline AMD severity group 2 or 3 and ≥ 1 year follow-up with complete data on macular phenotype, genetic data, dietary data including valid daily caloric intake [women: 600-3200 kcal and men: 600-4200 kcal] and family history of AMD, were divided into tertiles according to a genetic risk score (GRS) for progression. The weighted GRS was calculated using beta coefficients multiplied by the number of minor alleles present for 12 variants in 9 genes as provided in the **Supplemental Appendix.**^15^ This GRS was previously used in analyses of eyes transitioning to higher non-advanced stages of AMD or progressing to AAMD, adjusted for age, race and sex. ^15,16^ Genetic variants related to progression included *CFH* Y402H (rs1061170), *ARMS2/ HTRA1* A69S (rs10490924), *CFH* (rs1410996), *CFH* R1210C (rs121913059), *C3* R102G (rs2230199), *C3* K155Q (rs147859257), RAD51 paralog B (*RAD51B*) (rs8017304), transforming growth factor-beta receptor type 1 (*TGFBR1*) (rs334353), ATP-binding cassette transporter (*ABCA1*) (rs1883025), heat shock protein family H (Hsp110) member 1/beta 3-glucosyltransferase (*HSPH1/B3GALTL*) (rs9542236), tissue inhibitor of metalloproteinases 3 (*TIMP3*) (rs9621532), solute carrier family 16 member 8 (*SLC16A8*) (rs8135665). These variants are in the complement, immune/inflammatory, DNA repair, angiogenesis, lipid and extracellular matrix pathways. The protective variants were recoded such that each component of the GRS had a positive association with progression. The GRS was divided into tertiles and subjects in the highest tertile were defined as being at high genetic risk. We cross-classified GRS tertiles with baseline AMD severity groups and family history of AMD separately. Follow-up time was limited to a 5-year period to reduce misclassification of exposures related to lifestyle changes.

**Figure 1.**
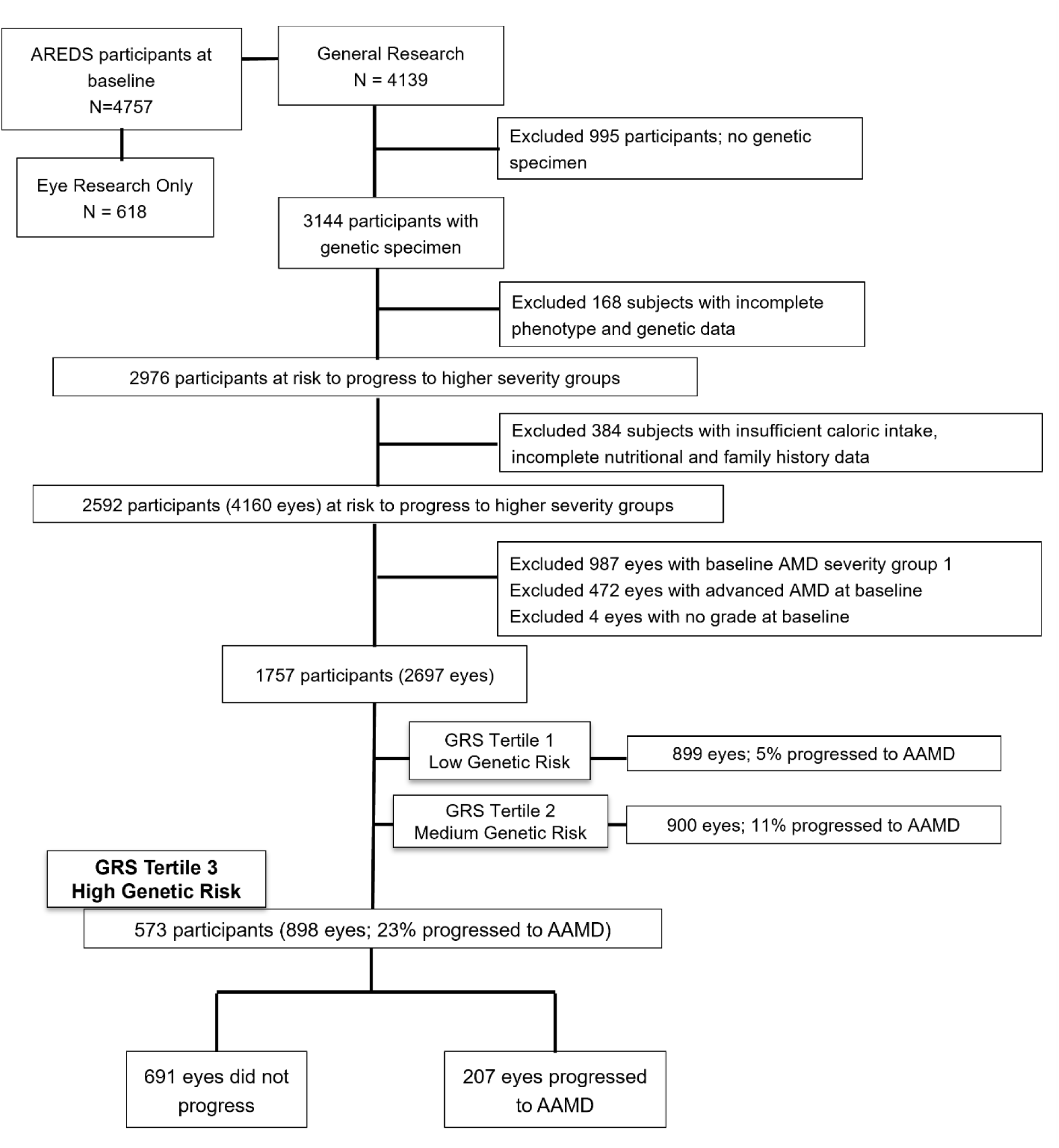
Flowchart showing selection of subjects for this analysis of subjects with high genetic risk (GRS tertile 3) for progression to AAMD from the AREDS cohort (*N* = 898 eyes). AMD, age-related macular degeneration; AAMD, advanced age-related macular degeneration; AREDS, Age-Related Eye Disease Study; GRS, genetic risk score.

### Measurement of exposure: risk-inducing vs. health-promoting behaviors

Previous studies have established modifiable lifestyle factors associated with progression to AAMD: smoking, BMI, daily caloric intake and a diet rich in green leafy vegetables and fish. ^3–8^ These variables were included in the analysis as follows:

i. Smoking status - Data on smoking status was collected at baseline as never, past or current smoker. Never smoking was considered an ideal health promoting behavior. Past and current smokers were combined as ever smokers to increase sample size within the comparison groups. The category of never smoking was considered to be a constant unchanging characteristic since few never smokers at baseline begin to smoke in late adulthood. On the other hand, ever smokers may change status from current to past and back to current, therefore as part of the lifestyle profile for ever smokers, current smoking is a risk-inducing behavior while past smoking is a health-promoting behavior.
ii. BMI (kg/m^2^)– BMI was calculated from height and weight measurements from baseline data. BMI was then dichotomized as ≥ 25 (i.e. a risk-inducing behavior) vs. < 25 (i.e. health-promoting behavior).
iii. Caloric Intake (kcal) – Daily caloric intake was obtained from the validated, self-administered, 90-item semi-quantitative AREDS food frequency questionnaire based on the National Cancer Institute Health Habits and History Questionnaire (v2.1) by the participant at a baseline visit. The University of Minnesota Nutrition Coordinating Center Food Composition Database (v31) was used to estimate quantity of caloric intake. The median caloric intake was 1701 kcal/day for males and 1261 kcal/day for females. Caloric intake was dichotomized into caloric intake ≥ sex specific median (i.e. risk-inducing behavior) and an intake of < sex-specific median (i.e. health-promoting behavior).
iv. Green Leafy Vegetables – Weekly ½ cup servings of green leafy vegetables rich in lutein-zeaxanthin were estimated using the AREDS food frequency questionnaire (e.g.: spinach (raw or cooked), greens (cooked), mustard greens, turnip greens, collards). Based on previous analysis, an average of ≥ 2.7 servings/week of green leafy vegetables was effective in reducing progression to both higher non-advanced and AAMD stages ^16^ and was considered a health-promoting behavior. A weekly intake of < 2.7 servings of green leafy vegetables was considered a risk-inducing behavior. This cut-off is similar to the USDA recommendations for elderly adults. ^23^
v. Fish – Weekly intake of fish rich in omega-3 fatty acids was also estimated (e.g. broiled or baked fish such as tuna, salmon, mackerel, trout). Fried fish was not included in the definition of fish intake. Fish intake was dichotomized as a weekly intake of < 2 medium servings (4 ounces) (i.e. risk-inducing behavior) and ≥ 2 medium servings/week (i.e. health-promoting behavior).

Lifestyle profiles were defined based on combinations of unhealthy and healthy behaviors. An ideal health-promoting lifestyle profile for never smokers was defined as adhering to each of the healthy behaviors (i.e. BMI < 25, caloric intake ≤ sex-specific median, ≥ 2.7 servings/week of green leafy vegetables and ≥ two 4 oz. servings of fish/week) compared to a high risk-inducing profile including all the unhealthy behaviors (i.e. BMI ≥ 25, caloric intake > sex-specific median, < 2.7 servings/week of green leafy vegetables and < two 4 oz. servings of fish/week). Ever smokers could reduce their risk by quitting smoking/not relapsing, in addition to adopting the other healthy behaviors listed above. The lifestyle profiles for ever and never smokers are detailed in the **Supplemental Appendix.**

### Other Covariates included in the analyses

Demographic variables included age groups (55-64, 65-74 and ≥75 years), sex (male/female) and education (≤ high school/>high school) obtained from baseline questionnaires. AREDS supplement group was defined as ‘Treatment’ if treated by antioxidants only, zinc only, or a combination of antioxidants and zinc and ‘Placebo’ for placebo assignment. Multivitamin intake was defined as ‘Yes’ for individuals who reported the intake of a multivitamin supplement at baseline, and ‘No’ for those with no self-reported intake. The baseline macular status of each eye was categorized into non-advanced AMD severity group 2 or group 3 and the fellow eye was categorized as non-advanced or advanced since an advanced fellow eye is a risk factor for progression of the other eye. ^14,24,25^ Family history of AMD (blood relatives including parents, siblings or cousins) was self-reported and classified as (a) not affected, (b) 1 family member affected with AMD or (c) ≥ 2 family members affected with AMD. This variable was included since it was independently associated with progression to AAMD, after adjusting for genetic variants. ^15^

### Main Outcomes

Progression was defined as an eye progressing from early or intermediate AMD to AAMD over 5 years. The outcome had to be confirmed across 2 consecutive visits occurring 6 months apart. ^15,16^ Time since baseline (quantified by the visit number) was used as the time scale for this analysis. An eye was censored at the earliest confirmed diagnosis of AAMD, at 5 years, loss to follow up or death. Further analyses evaluated progression to AAMD subtypes – GA and NV. There were some eyes which progressed to GA at 2 consecutive visits and subsequently progressed to NV at 2 consecutive visits; therefore, these eyes were counted as progressing to both GA and NV. However, if an eye progressed to NV and then to GA, follow-up was stopped at time of NV confirmation and no further visits were evaluated since some forms of GA could be related to treatment and complications of NV.

### Statistical Analyses

We used the PROC PHREG procedure in SAS (v9.4) to perform Cox proportional hazards regression survival analysis with the aggregate option to account for inter-eye correlation, since the eye was the unit of analysis for eye-specific outcomes. All statistical tests were performed as 2-tailed with an α level of 0.05.

Baseline characteristics of eyes that progressed versus eyes that did not progress were compared using univariate analyses, adjusting for age groups and baseline AMD severity groups. We further assessed a multivariate model (incorporating age, sex, education, AREDS supplement group, multivitamin intake, baseline macular phenotype, family history of AMD and unhealthy habits including smoking, BMI and diet) and defined a composite risk score computed as the sum of β coefficients for all variables as detailed in the **Supplemental Appendix**.

We aimed to determine the degree to which modifiable factors could impact disease risk among those with the highest genetic susceptibility. Using the ESTIMATE statement in PROC PHREG, we calculated Hazard Ratios (HRs) and 95% confidence intervals (CIs) to estimate the incidence of progression to AAMD over 5 years related to a risk-inducing profile as compared to a health-promoting profile in those with high genetic risk, adjusting for age group, sex, education, AREDS supplement group, multivitamin intake, fellow eye status, baseline AMD severity group and family history of AMD. Similar analyses were performed for progression to GA and NV.

Population attributable risk (PARs) further quantified the percentage of incident AAMD cases that could be prevented if eyes/individuals following an unhealthy lifestyle profile would adhere to an ideal health-promoting profile, assuming that the risk factors had no interactions. The PAR was calculated for various lifestyle profiles (*i*) created from 16 combinations of the behaviors for never smokers and 32 combinations for ever smokers, as given in the following formula:

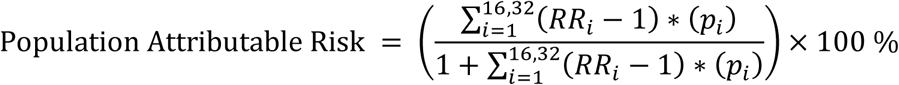

Relative risks (RRs) for associations between each lifestyle profile and progression to AAMD were estimated using linear combinations of the beta estimates from the multivariate adjusted model. These RRs were then combined with the prevalence (*p_i_*) of the specific lifestyle profiles to estimate the PAR.

We plotted direct adjusted cumulative incidence curves associated with risk of AMD progression based on the number of unhealthy behaviors, according to smoking status. Among never smokers with high genetic susceptibility, curves were plotted for low (0/1 out of 4), medium to high (2/3/4 out of 4) number of unhealthy behaviors. Similarly, among ever smokers, curves were plotted for low (0/1 out of 5), medium (2/3 out of 5) and high (4/5 out of 5) number of unhealthy habits.

We used the multivariate model (incorporating age, sex, education, AREDS supplement group, multivitamin intake, baseline macular phenotype, family history of AMD and unhealthy habits including smoking, BMI and diet), to estimate the cumulative 5-year incidence of AAMD, adjusted for competing mortality risks. Finally, for a few representative eyes selected from this cohort, we used baseline behaviors provided in the questionnaires and depicted the estimated impact of modifying certain lifestyle behaviors on their risk for progressing to AAMD using risk categories of low (0 to <10%), medium (10 to <30%), high (30 to < 50%) and very high (≥ 50%) risk, as previously described. ^15,26^

## RESULTS

As shown in **Table 1**, we divided the cohort into GRS tertiles. There were 898 eyes from 573 participants in the high genetic risk group (i.e. tertile 3) and among these, 207 eyes (23%) progressed to AAMD which was twice the progression rate in tertile 2 (11%) and four times the rate in tertile 1 (5%). The rate of progression to AAMD was higher in the baseline intermediate AMD severity group compared to eyes with early AMD within each genetic tertile (GRS tertile 3: 32% vs 7%, tertile 2: 20% vs. 2% and tertile 1: 16% vs 1%). Among the 207 progressing eyes in tertile 3, 93 developed GA and 118 developed NV (4 eyes developed GA and then subsequently progressed to NV). Similar differences in progression rates according to GRS and baseline AMD severity group were seen for progression to the advanced subtypes (**Supplemental Tables 1 and 2**). In addition, the higher GRS tertile was associated with having a family history of AMD (**Supplemental Table 3**). The percentage of people with ≥ 2 family members affected with AMD was 51% for tertile 3, 30% for tertile 2 and 19% for tertile 1 in comparison to 28%, 34% and 38%, respectively, for those with no affected family members. The odds of being in GRS tertile 3 vs. tertile 1 for subjects with ≥ 2 family members affected was 3.6 times higher than the odds of GRS tertile 3 vs. tertile 1 for subjects with no family members (Odds Ratio = 3.6, 95% CI 2.3-5.6, P<0.001). The remainder of the analyses focuses on eyes in the highest genetic tertile.

**Table 1.** Association of Progression to Advanced AMD by Baseline AMD severity group and GRS tertile over 5 years.

|  |  |  |  |  |  |  |  |  |
| --- | --- | --- | --- | --- | --- | --- | --- | --- |
|  |  | GRS Tertiles |  |  |  |  |  | Total eyes |
|  |  | Tertile 1 (Low Genetic Risk)<br>0.263-1.458 |  | Tertile 2 (Medium Genetic Risk)<br>1.459-1.881 |  | Tertile 3 (High Genetic Risk)<br>1.882-3.108 |  |  |
|  |  | Total | Progressing eyes<br>(Progression Rate to<br>AAMD) | Total | Progressing eyes,<br>(Progression Rate to<br>AAMD) | Total | Progressing eyes,<br>(Progression Rate to<br>AAMD) |  |
| Baseline<br>AMD<br>Severity<br>Group | Group 2 (early AMD) | 633 | 6 (1%) | 463 | 9 (2%) | 309 | 21 (7%) | 1405 |
|  | Group 3 (intermediate AMD) | 266 | 42 (16%) | 437 | 88 (20%) | 589 | 186 (32%) | 1292 |
| Total eyes |  | 899 | 48 (5%) | 900 | 97 (11%) | 898 | 207 (23%) | 2697 |

AMD = age related macular degeneration; AAMD = advanced age related macular degeneration; GRS = genetic risk score

Associations between progression to AAMD and covariates are shown in **Table 2**. In eyes with high genetic risk, adjusting for age groups and baseline AMD severity (Model 1), increased incidence of progression was significantly associated with older age, fellow eyes being advanced at baseline, and eyes with AMD severity group 3 vs. group 2 at baseline. Increased incidence of progression was also associated with current smoking, high BMI, and fish intake < 2 medium servings/week. In the multivariate model adjusted for all covariates (Model 2), similar effect sizes were seen for BMI and fish intake while non-significant trends for increased risk of progression were seen with current smoking, high caloric intake, low intake of green leafy vegetables, as well as more family members affected with AMD. In this high genetic risk population, the composite risk score computed from the estimates of this multivariable model, ranked by quartiles, was plotted for all eyes in **Figure 2** by progression status. As the score increased, there was a higher proportion of eyes that progressed to AAMD (HR for quartile 4 vs quartile 1 = 13.8, 95% CI =7.3-26.1, P<0.001; P_trend_<0.001).

**Figure 2.**
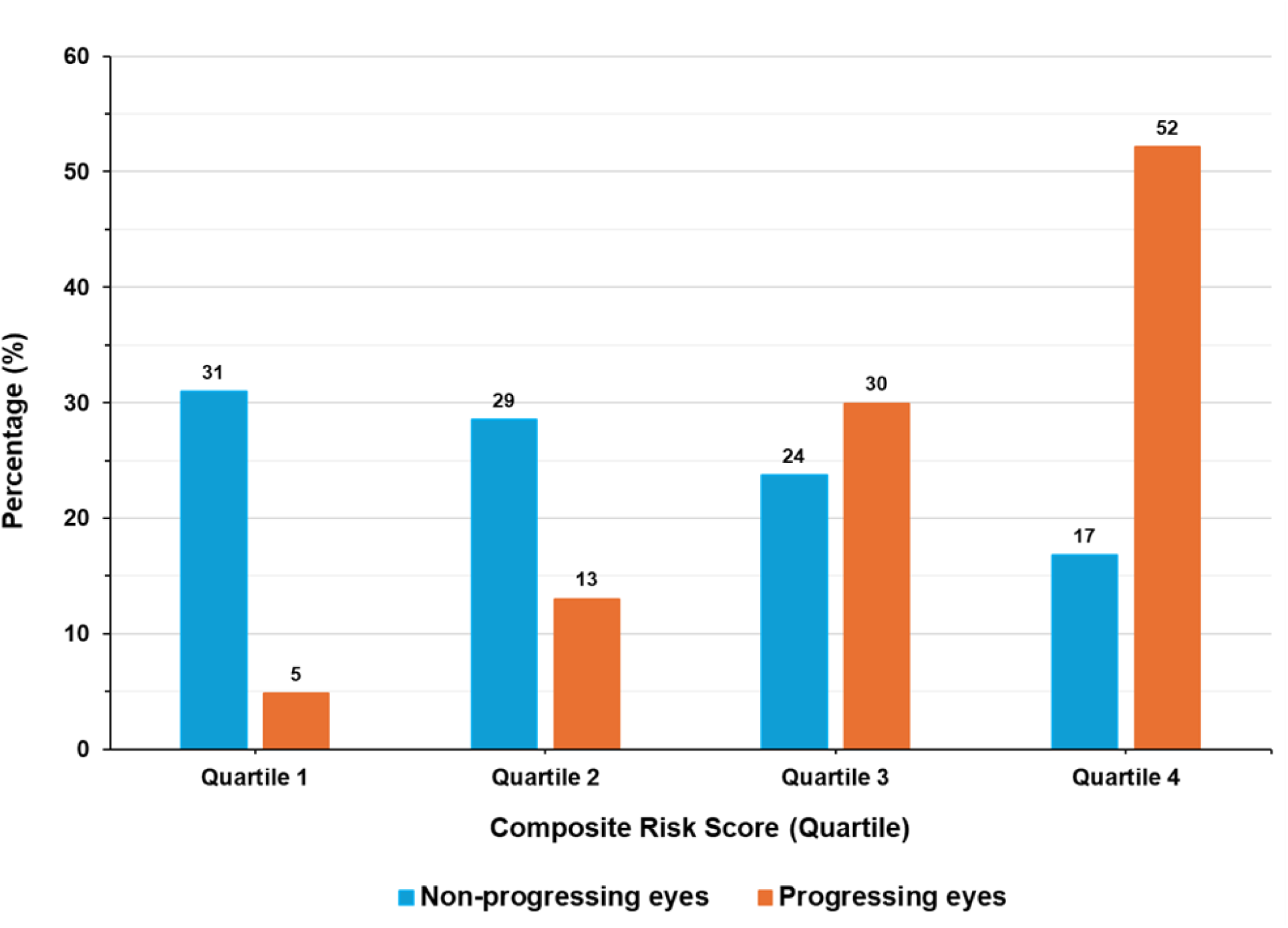
Distribution of genetically high-risk eyes by progression status according to the composite risk score (quartiles) which was computed using beta estimates from Table 2, Model 2. Hazard Ratio for quartile 4 vs quartile 1 = 13.8, 95% CI =7.3-26.1, P<0.001; P_trend_<0.001

**Table 2.**
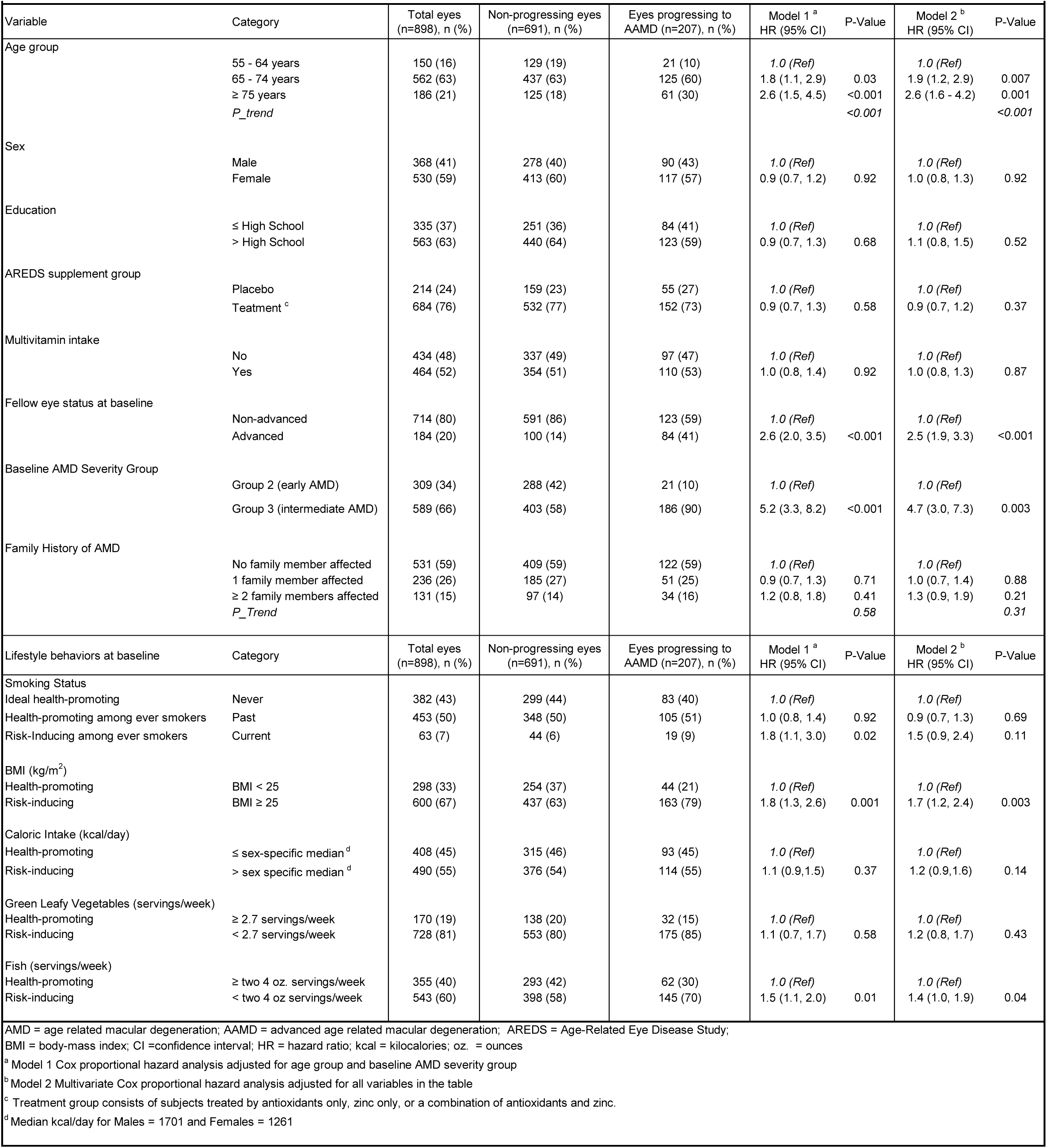
Associations between progression to AAMD over 5 years and covariates in eyes with high genetic risk (N = 207 events/898 eyes)

The associations between a high risk-inducing lifestyle profile and progression to AAMD over 5 years compared to an ideal health-promoting profile are shown in **Table 3**. In never smokers, the high-risk inducing profile conferred over a 3-fold increased incidence of progression to AAMD (HR = 3.3, CI 1.8-6.4, P<0.001), and over a 5-fold higher incidence of progression in those who had ever smoked (HR = 5.3, CI 2.3-11.9, P<0.001). The AUC of the predictive model with all covariates was 0.77 for this high genetic risk population. Analyses of these profiles and progression rates to AAMD were evaluated separately in the AREDS supplement treatment and placebo groups and similar results were seen (**Supplemental Tables 4 and 5**). There was a significant adverse effect of risk-inducing vs. ideal health promoting lifestyles in both groups for never and ever smokers. The consistency of these findings for supplement users and those taking placebo suggests that a health-promoting lifestyle plays a fundamental role in reducing progression to AAMD, which may not be mitigated by nutritional supplementation alone in this high genetic risk group.

**Table 3.** Association between progression to AAMD over 5 years and risk-inducing profile at baseline among eyes with high genetic risk according to smoking status N= 207 events/898 eyes.

| Lifestyle Profile | Never Smokers<br>n=83 events/382 eyes |  | Ever Smokers<br>n=124 events/516 eyes |  |
| --- | --- | --- | --- | --- |
|  | HR (95% CI) <sup>c</sup> | P-value | HR (95% CI) <sup>c</sup> | P-value |
| Ideal health-promoting profile <sup>a</sup> | 1.0 (Ref) | - | 1.0 (Ref) | - |
| High risk-inducing profile <sup>b</sup> | 3.3 (1.8, 6.4) | <0.001 | 5.3 (2.3, 11.9) | <0.001 |

AAMD = advanced age-related macular degeneration; AMD = Age-related macular degeneration
AREDS = Age-Related Eye Disease Study
<sup>a</sup> A health-promoting profile for never smokers consists of BMI < 25, caloric intake/day ≤ sex-specific median, green leafy vegetables ≥ 2.7 servings/week and fish intake ≥ 2 servings/week; for ever smokers the profile also includes past smoking
<sup>b</sup> A risk-inducing profile for never smokers consists of BMI ≥ 25, caloric intake/day > sex-specific median, green leafy vegetables < 2.7 servings/ week and fish intake < 2 servings/week; for ever smokers the profile also includes current smoking
<sup>c</sup> HRs estimated using linear combination of the beta estimates from Table 2, model 2 adjusted for age group, sex, education, AREDS supplement group, multivitamin intake, fellow eye status, baseline AMD severity group and family history of AMD

Associations between the lifestyle profiles and incidence of progression to advanced GA and NV subtypes are shown in **Tables 4 and 5**. For both never and ever smokers, the deleterious effects of the high risk-inducing lifestyle compared to an ideal health-promoting lifestyle was more evident for GA (P< 0.001) than NV (P = 0.09 to 0.23), although effects were in the same directions for both. Multivariate estimates for individual lifestyle factors for AAMD, GA and NV are provided in **Supplemental Figure 1.** The hazard ratio for current smoking as a risk factor for progression was higher for NV than GA, whereas each of the hazard ratios for the effects of high BMI, high caloric intake, low intake of green leafy vegetables and fish on progression to advanced stages were higher for GA than NV.

**Table 4.** Association between progression to GA over 5 years and risk-inducing profile at baseline among eyes with high genetic risk according to smoking status N= 93 events/898 eyes.

|  |  |  |  |  |
| --- | --- | --- | --- | --- |
| Lifestyle Profile | Never Smokers<br>(n= 41 events/382 eyes) |  | Ever Smokers<br>(n= 52 events/516 eyes) |  |
|  | HR (95% CI) <sup>c</sup> | P-value | HR (95% CI) <sup>c</sup> | P-value |
| Ideal health-promoting profile <sup>a</sup> | 1.0 (*Ref*) | - | 1.0 (*Ref*) | - |
| High risk-inducing profile <sup>b</sup> | 8.1 (3.1, 21.1) | <0.001 | 11.9 (3.4, 42.3) | <0.001 |

AMD = age-related macular degeneration; AREDS = Age-Related Eye Disease Study
CI = Confidence Interval; GA = geographic atrophy; HR = Hazard Ratio
<sup>a</sup> A health-promoting profile for never smokers consists of BMI < 25, caloric intake/day ≤ sex-specific median, green leafy vegetables ≥ 2.7 servings/week and fish intake ≥ 2 servings/week; for ever smokers the profile also includes past smoking
<sup>b</sup> A risk-inducing profile for never smokers consists of BMI ≥ 25, caloric intake/day > sex-specific median, green leafy vegetables < 2.7 servings/ week and fish intake < 2 servings/week; for ever smokers the profile also includes current smoking
<sup>c</sup> HRs estimated using linear combination of the beta estimates from Table 2, model 2 adjusted for age group, sex, education, AREDS treatment group, multivitamin intake, fellow eye status, baseline AMD severity group and family history of AMD

**Table 5.** Association between progression to NV over 5 years and risk-inducing profile at baseline among eyes with high genetic risk according to smoking status N= 118 events/898 eyes.

| **N= 118 events/898 eyes** |  |  |  |  |
| --- | --- | --- | --- | --- |
| Lifestyle Profile | Never Smokers<br>(n=43 events/382 eyes) |  | Ever Smokers<br>(n= 75 events/516 eyes) |  |
|  | HR (95% CI) <sup>c</sup> | P-value | HR (95% CI) <sup>c</sup> | P-value |
| Ideal health-promoting profile <sup>a</sup> | 1.0 (*Ref*) | - | 1.0 (*Ref*) | - |
| High risk-inducing profile <sup>b</sup> | 1.7 (0.7, 4.0) | 0.23 | 2.6 (0.8, 7.7) | 0.09 |

AMD = age-related macular degeneration; AREDS = Age-Related Eye Disease Study; CI = Confidence Interval; HR = Hazard Ratio; NV = neovascular
<sup>a</sup> A health-promoting profile for never smokers consists of BMI < 25, caloric intake/day ≤ sex-specific median, green leafy vegetables ≥ 2.7 servings/week and fish intake ≥ 2 servings/week; for ever smokers the profile also includes past smoking
<sup>b</sup> A risk-inducing profile for never smokers consists of BMI ≥ 25, caloric intake/day > sex-specific median, green leafy vegetables < 2.7 servings/ week and fish intake < 2 servings/week; for ever smokers the profile also includes current smoking
<sup>c</sup> HRs estimated using linear combination of the beta estimates from Table 2, model 2 adjusted for age group, sex, education, AREDS treatment group, multivitamin intake, fellow eye status, baseline AMD severity group and family history of AMD

The RRs of the different lifestyle profiles and the number of unhealthy behaviors for never and ever smokers are shown in **Supplementary Tables 6 and 7**. An ideal health-promoting profile was considered the reference. As the number of unhealthy behaviors increased, there was a gradual increase in the RRs for progression ranging from 1.2 to 3.3 for never smokers and 1.2 to 5.3 for ever smokers. The PARs for never smokers and ever smokers for the modifiable risk factors were 56% and 60% respectively, after adjusting for covariates. These results indicate that if everyone in the study population who followed one or more unhealthy behaviors adopted the corresponding healthy behaviors, the incidence of AAMD would be reduced by 56% for never smokers and 60% for ever smokers.

**Figure 3** displays the RRs of progressing to AAMD by the number of unhealthy risk-inducing behavior(s) as compared to the reference, according to smoking status. As the number of unhealthy behaviors increased, the RR increased in both never and ever smokers. Smokers had a higher RR for more than 3 unhealthy behaviors than never smokers. These results indicate the significant impact of modifying unhealthy lifestyle behaviors in a high genetic risk population to prevent the progression of early and intermediate disease to the advanced stage over 5 years.

**Figure 3.**
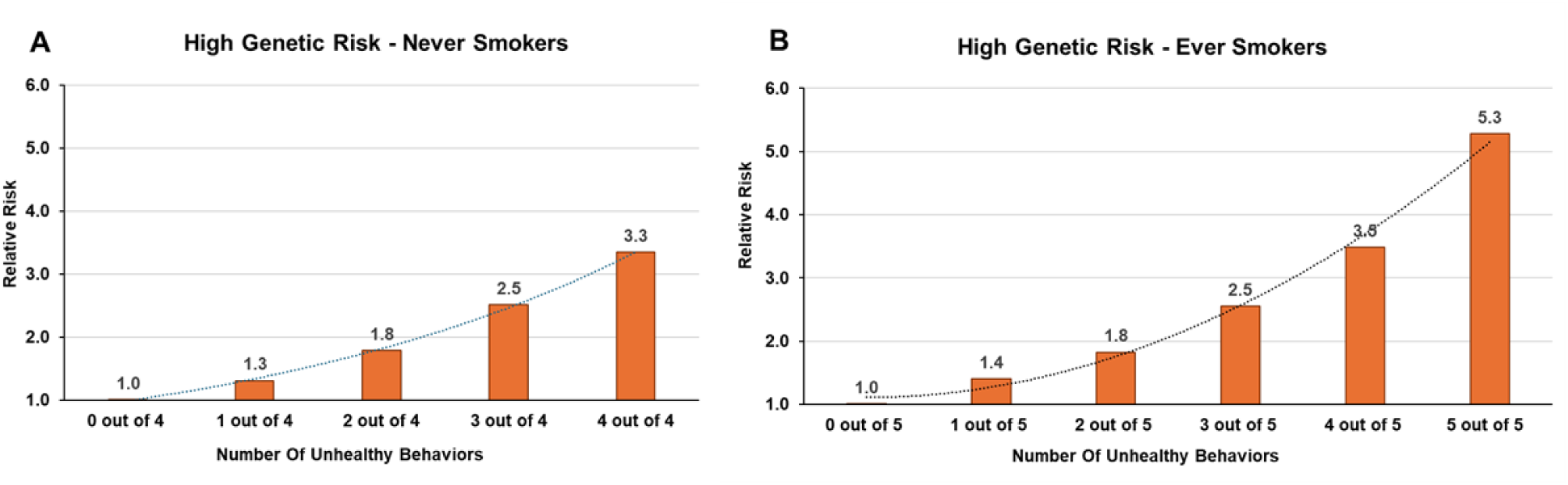
Relative risk of progressing to advanced age-related macular degeneration associated with the number of unhealthy risk-inducing behaviors compared to an ideal health-promoting profile among A) never smokers and B) ever smokers with high genetic risk. Risk inducing behaviors include high body-mass index (BMI) (≥ 25), daily caloric intake > sex-specific median, intake of green leafy vegetables < 2.7 serving/week, fish intake < 2 medium servings/week compared to an ideal health-promoting lifestyle of low BMI (<25), daily caloric intake ≤ sex-specific median, intake of green leafy vegetables ≥ 2.7 serving/week, fish intake ≥ 2 medium servings/week. In ever smokers, the profile also includes current smoking as a risk-inducing behavior compared to a past smoker.

Direct adjusted cumulative incidence curves by number of unhealthy behaviors in never and ever smokers with high genetic risk are displayed in **Figures 4 and 5**. Over the 5-year follow-up period, unhealthy behaviors were associated with an overall increased risk of eyes progressing to AAMD. The cumulative incidence of progression to AAMD increased to 23% for never smokers with medium to high number of unhealthy behaviors (**Figure 4**). In ever smokers (**Figure 5**), the cumulative incidence was higher and increased from 17% to 20% to 31% with an increasing number of unhealthy behaviors. With optimal lifestyle habits, the cumulative incidence of AAMD over 5 years was lower, but was still 5% and 17% in never and ever smokers, likely due to the underlying high genetic risk of this study population. As a check on the fit of the model, we computed the actual percentage of eyes that progressed to AAMD over 5 years among never smokers with a specific number of unhealthy behaviors. We found that for 66 eyes from subjects who never smoked and had all 4 unhealthy behaviors, 18 eyes progressed to AAMD (27%), compared with 51 eyes from subjects who never smoked and had only 0/1 unhealthy behaviors, among whom 2 eyes progressed to AAMD (2%) over 5 years, which corresponds closely to the estimates in **Figure 4**.

**Figure 4.**
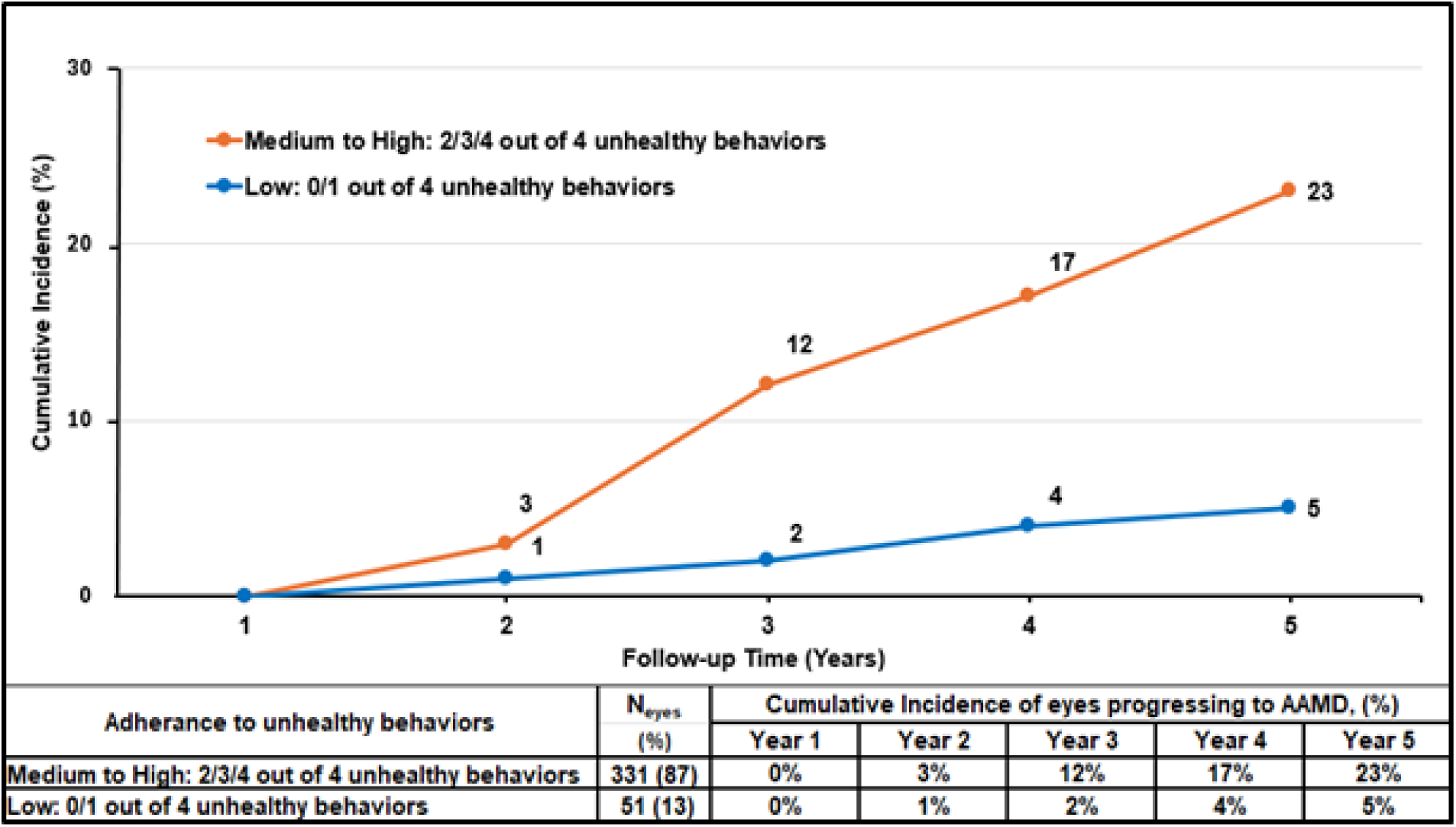
Cumulative incidence curves for AAMD among never smokers with high genetic risk by number of unhealthy behaviors, direct adjusted for age group, sex, education, AREDS supplement group, multivitamin intake, fellow eye status, baseline AMD severity group and family history of AMD. Unhealthy behaviors included body-mass index ≥ 25, caloric intake > sex-specific median, < 2.7 servings/week of green leafy vegetables and < two 4 oz. servings of fish/week. AMD, age-related macular degeneration; AAMD, advanced AMD; AREDS, Age-Related Eye Disease Study

**Figure 5.**
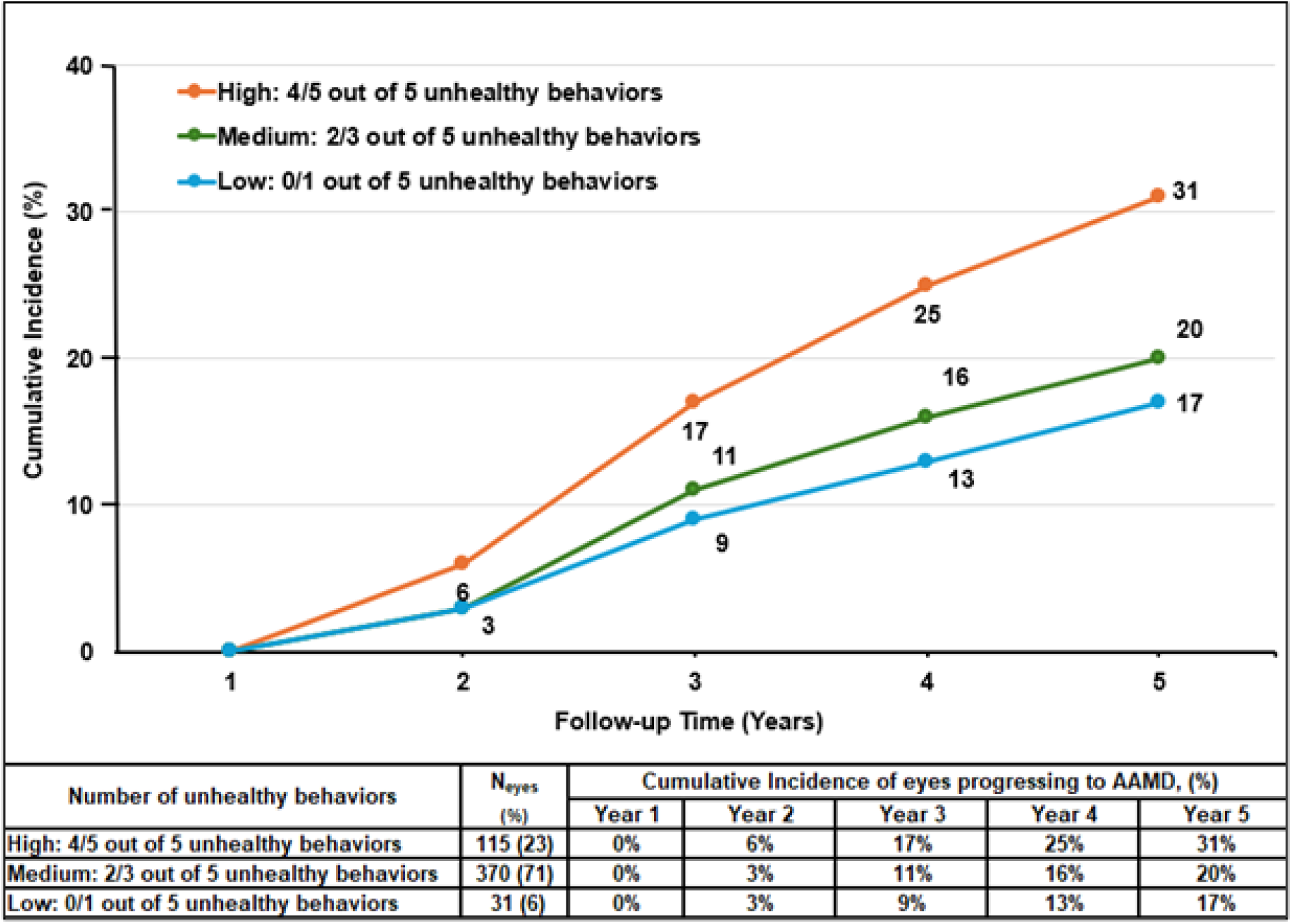
Cumulative incidence curves for AAMD among ever smokers with high genetic risk by number of unhealthy behaviors, direct adjusted for age group, sex, education, AREDS supplement group, multivitamin intake, fellow eye status, baseline AMD severity group and family history of AMD. Unhealthy behaviors included smoking, body-mass index ≥ 25, caloric intake > sex-specific median, < 2.7 servings/week of green leafy vegetables and < two 4 oz. servings of fish/week. AMD, age-related macular degeneration; AAMD, advanced AMD; AREDS, Age-Related Eye Disease Study

We now consider the predicted 5-year risk of progression to AAMD for three genetically high-risk individuals (actual age hidden for deidentification), adjusted for competing mortality risks, shown in **Figure 6**.

**Figure 6.**
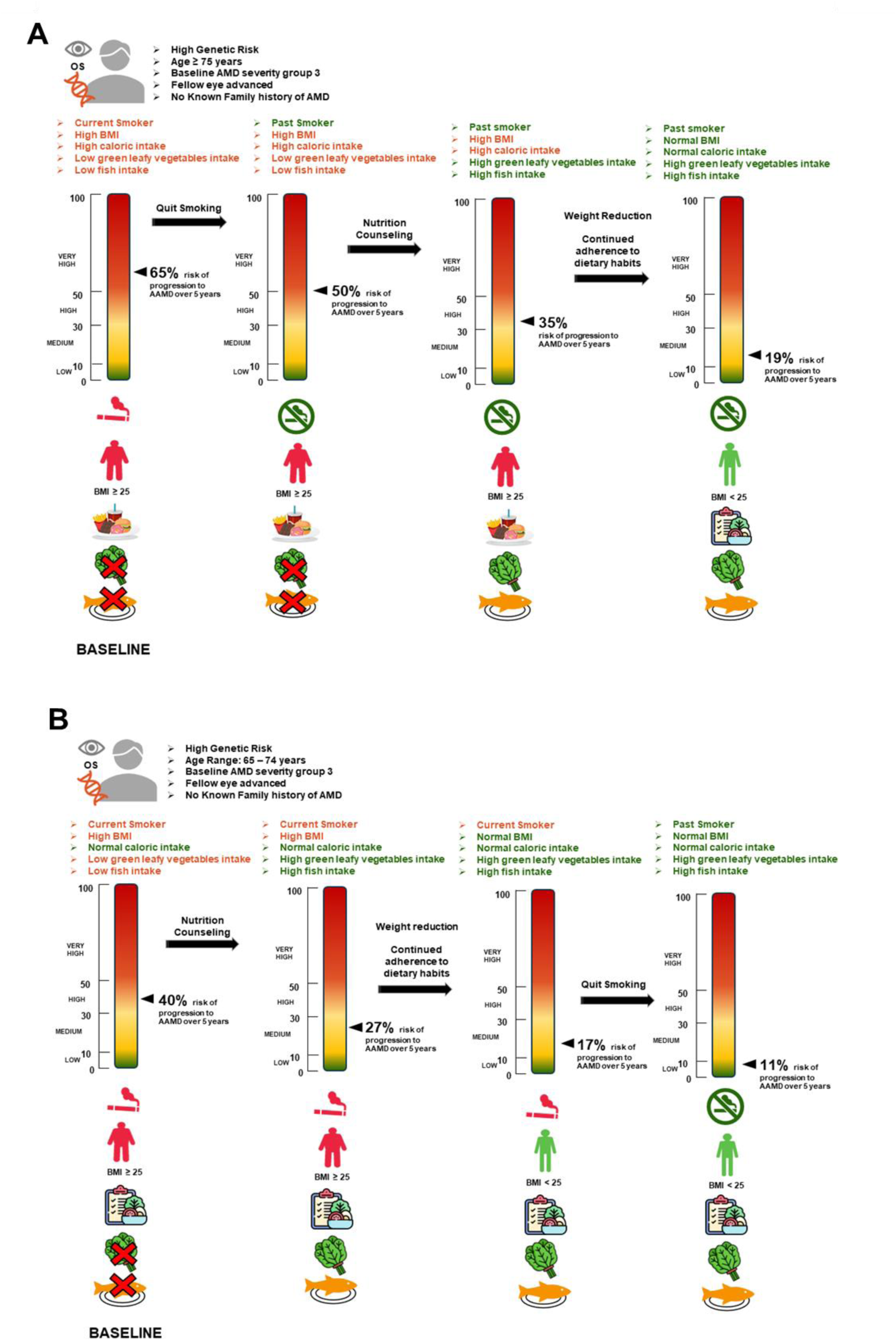
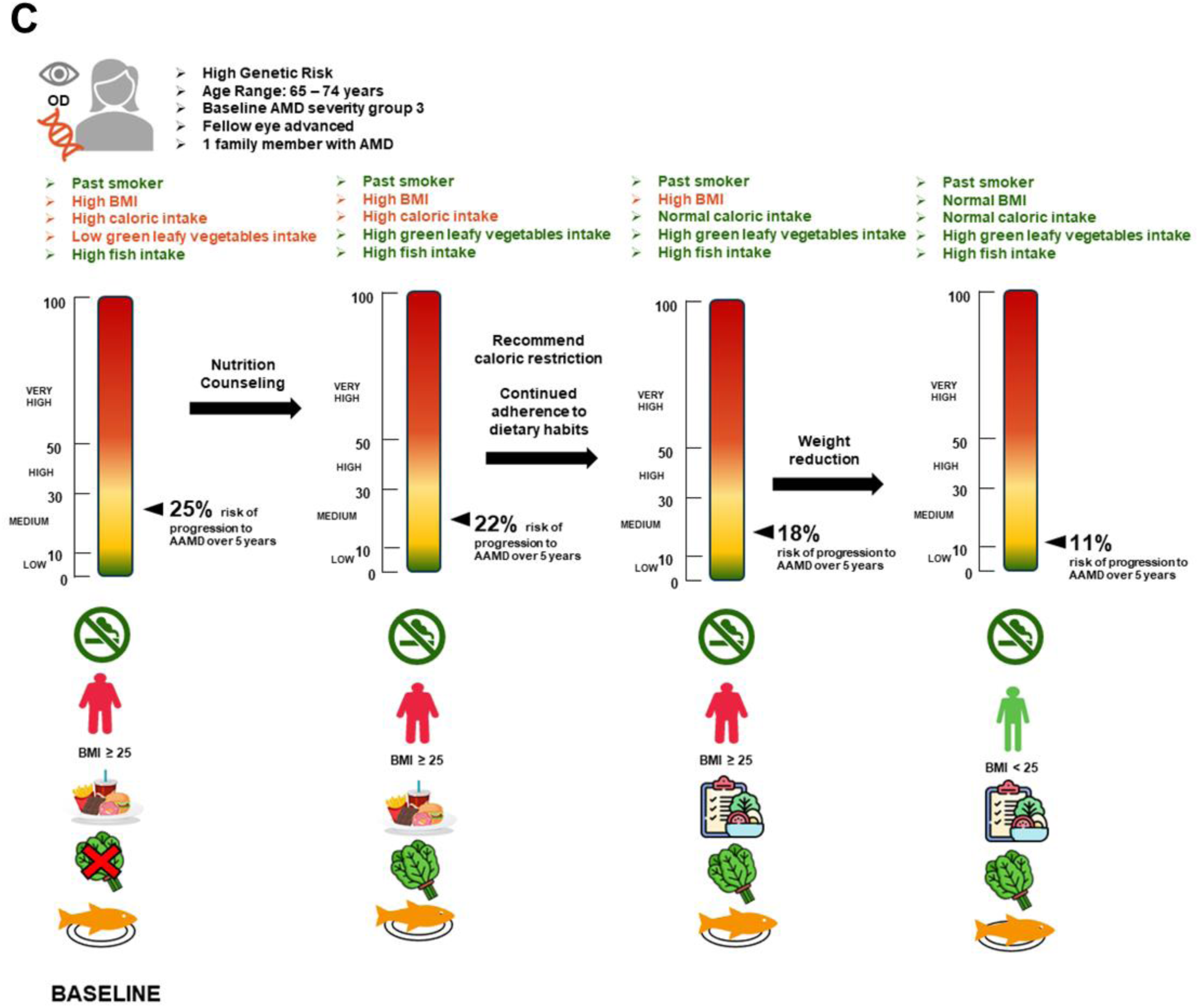
<u>Case A:</u> Male aged ≥ 75 years old with intermediate AMD OS and advanced fellow eye at baseline followed a high risk-inducing lifestyle (5 out of 5 unhealthy behaviors). With the adoption of healthy lifestyle behaviors, the risk for progression to AAMD is projected to decrease from 65% to 50% to 35% to 19%. <u>Case B</u>: Male aged between 65-74 years with intermediate AMD OS and advanced fellow eye at baseline followed 4 out of 5 unhealthy behaviors but had a normal caloric intake. The risk for progression to AAMD is projected to decrease from 40% to 27% to 17% to 11% with the reduction in number of unhealthy behaviors and developing an ideal health-promoting lifestyle. <u>Case C</u>: Female aged between 65-74 years, with intermediate AMD OD and advanced AMD in the fellow eye at baseline. She had 1 family member affected with AMD. She was a past smoker with adequate fish intake and had 3 out of 5 other unhealthy behaviors. Adherence to an ideal health-promoting lifestyle is projected to decrease AAMD progression from 25% to 11%. AMD, age-related macular degeneration; AAMD, advanced AMD

Case A was a male aged ≥ 75 years, current smoker, with intermediate AMD OS and advanced disease in the fellow eye at baseline. He had no known family history of AMD and followed a risk-inducing lifestyle (5 out of 5 unhealthy behaviors). If he quit smoking, the risk was projected to decrease from 65% to 50%. With nutrition counseling and weight reduction, the risk for AAMD was projected to decrease to 19%.

Case B was a male aged between 65-74 years, current smoker, with intermediate AMD OS and advanced AMD in the fellow eye at baseline. He had no known family history of AMD and followed 4 out of 5 unhealthy behaviors but had a normal caloric intake. With nutrition counseling to incorporate green leafy vegetables and fish in his weekly diet, AAMD progression was projected to decrease from 40% to 27%. Furthermore, BMI reduction to < 25 together with dietary adherence would decrease AAMD incidence to 17%. Quitting smoking plus adherence to other healthy habits would decrease the risk of AAMD progression to 11% over 5 years.

Case C was a female aged between 65-74 years, past smoker with adequate fish intake, with intermediate AMD OD and advanced AMD in the fellow eye at baseline. She had 1 family member affected with AMD and reported 3 out of 5 unhealthy behaviors. With nutrition counseling to increase green leafy consumption, caloric restriction and weight reduction, progression to AAMD was projected to decrease from 25% to 11% over 5 years.

These case examples underscore the significant impact of multiple unhealthy behaviors on AAMD progression over time, particularly in this genetically susceptible population, emphasizing the critical role of lifestyle factors in managing AMD risk.

## DISCUSSION

### Main Results

In this study we have quantified the effects of an unhealthy lifestyle profile on progression from non-advanced AMD to AAMD over 5 years among genetically high-risk eyes. A risk-inducing lifestyle raised the likelihood of progression to AAMD by 3-fold in never smokers and over 5-fold among smokers in this genetically susceptible population. After accounting for a high genetic risk and other covariates, 56-60% of AAMD incidence was attributed to the modifiable risk factors considered in this study.

Analyses of the advanced AMD subtypes showed that a risk inducing lifestyle was associated with an 8-to-12-fold higher risk of progression to GA, and a lower but relevant 2-to-3-fold increased risk of progression to NV, in never and ever smokers respectively. Consistent with results in this high genetic risk cohort, associations between high BMI and progression to GA, and a greater effect of smoking on incidence of NV have been reported for AMD overall. ^14,15,24–27^ Differences in the association between lifestyle behaviors and the advanced subtypes may reflect pathogenic variations in oxidative stress, dysregulations in inflammatory/immune or lipid pathways, or other differential biologic effects on the retinal pigment epithelium, outer retina, Bruch’s membrane and/or choroid. Lifestyle profiles play a key role in progression to both GA and NV, and high genetic susceptibility in this cohort may attenuate the effects of behaviors on progression to NV. These comparisons between GA and NV in individuals with high genetic burden require further investigation.

The case examples in this study further highlight the clinical application of personalized lifestyle modifications to reduce the 5-year risk of progression to AAMD, even among individuals with a high genetic risk. While family history of AMD is associated with genetic risk, our results indicate that a positive family history is not a proxy for high GRS and vice versa. Even with no family history of the disease, 28% of the population had a baseline vulnerability with a high GRS, and behavioral modifications could offer significant opportunities for intervention.

These results emphasize the importance of comprehensive motivational lifestyle counseling including smoking cessation, maintaining a healthy weight, avoiding high daily caloric intake and incorporating foods rich in lutein-zeaxanthin (e.g.: spinach, kale, collards, mustard greens, turnip greens, brussels sprouts, broccoli) and fatty fishes rich in omega-3 fatty acids (e.g.: salmon, sardines, mackerel, tuna, and trout). Encouraging personalized lifestyle modifications over multiple clinical visits may be a key strategy to reduce AAMD progression over time and to improve patient outcomes, even in those with increased genetic burden who are more likely to progress. Individuals with low or medium genetic risk should also be encouraged to adhere to a health-promoting lifestyle.

### Mechanism of gene-lifestyle associations

Our previous work has demonstrated that the GRS, consisting of selected genetic variants related to progression, has a strong positive association with progression to AAMD, GA, NV ^14,15,24,27^ and transitions to higher non-advanced AMD severity stages.^16^ However, underscoring that genetics do not act alone in this complex disease, twin studies of monozygotic twins discordant with respect to their AMD diagnosis have demonstrated association of higher unhealthy behaviors in the twin with more advanced disease ^7,26^ Behavioral lifestyles could result in epigenetic modifications through DNA methylation and subsequent altered gene expression in AMD. ^29^ These include protective behavioral factors such as lower abdominal adiposity and BMI ^6,27,30^, not smoking ^4,7,27,28,30^ and dietary intake of foods rich in vitamin D, methionine, betaine, lutein/zeaxanthin, omega-3 fatty acid, thiamin, riboflavin, and folate which have been reported to reduce the overall risk of AMD onset ^3,7,28,31^ and progression. ^10,12,16,32^ The bioactive components in green leafy vegetables and fish including the carotenoids lutein and zeaxanthin, and omega-3 fatty acids, have been associated with protective effects on development and progression of AMD. ^3,7,10,11,16,33^ Potential mechanisms include the effects of these foods and nutrients on reducing oxidative stress and inflammation, maintenance of retinal pigment epithelium integrity, and protection against photo-oxidative damage to the macula.^34–36^ Polymorphisms in the most frequently studied *CFH* (complement/immune pathway) and *ARMS2/HTRA1* (potentially inflammatory pathway) genes ^37^ and variants in our GRS have also been independently linked to progression of the disease. ^2,15,24,27,38,39^ Adoption of a sustainable healthy lifestyle in the high GRS group could lead to a reduction in complement activation, oxidative stress, systemic inflammation, serum levels of C-reactive protein,^34,40^ and overall decreased incidence of AAMD. Recent research has also shed light on the gut microbiome-retina connection to AMD which further cements the importance of diet and genetics in relation to AMD progression.^41^ This interplay underscores the importance of considering both genetic and environmental factors in understanding the mechanisms behind AMD progression and promoting an integrated approach towards preventing visual impairment in those with high genetic susceptibility.

### Other Reports

Previous studies have examined the interactions between genetic susceptibility and lifestyle factors in relation to AMD risk. A 2006 case-control analysis reported strong susceptibility of the joint effects of the high risk *CFH* Y402H (CC genotype) plus a higher BMI (OR = 5.9) or high risk genotype plus smoking (OR = 10.2) on advanced AMD. ^30^ A 2013 study reported increased intake of DHA and reduced progression to GA among those with *the ARMS2/HTRA1* homozygous risk genotype (HR=0.4; P = 0.002) or those with *CFH* Y402H homozygous non-risk genotype (HR = 0.5; P = 0.02). ^11^ The first study to analyze adherence to a Mediterranean diet and a genetic risk score on progression to AAMD reported a lower risk of incident AAMD among subjects carrying the *CFH* Y402H non-risk (T) allele (P-trend = 0.0004, P-interaction = 0.04) and associations of a higher dietary score with a high genetic risk score (HR = 0.75, P-value = 0.01). ^10^ A beneficial effect of the Mediterranean diet and gene interactions were also evaluated in other studies. ^18,42^

The effects of genes and lifestyles on prevalence of AMD using cross-sectional data has been assessed in various population based cohorts. ^17,18^ The EYE-RISK Consortium ^17^ conducted a pooled analyses of cross-sectional data and evaluated associations between a lifestyle score, polygenic risk score and AMD status in subjects with intermediate to AAMD compared to early or no AMD. They concluded that adherence to a healthy lifestyle was associated with reduced prevalence of AMD across all genetic risk strata, with the strongest effect seen in individuals at high genetic risk. Limitations of this study included the cross-sectional design with no time-to-event component and therefore no information on disease progression over time. Second, the outcome was defined broadly as the presence of intermediate or advanced AMD, rather than progression between specific disease stages. Third, genetic risk variants used were based on prevalence and not progression. ^15,27,43^ Finally, the analysis was conducted at the person level, excluding controlling for eye-specific factors such as baseline AMD severity and fellow eye status, both of which are known predictors of progression and may vary between eyes within the same individual. Evaluation of incidence of AMD (defined in various ways) related to diet or lifestyles has also been reported for levels of genetic risk defined by 2 genes in population based _studies.19,20,31,44_

### Strengths and Limitations

Key strengths and differences in this study include 1) a comprehensive longitudinal prospective analysis of smoking, BMI, caloric intake, dietary intake of leafy greens and fish on progression to AAMD, GA and NV from earlier non-advanced stages, exclusively in a high genetic risk group, 2) a focus on the highest genetic risk tertile to address the specific question which arises in real world clinical care-“am I destined to develop AMD if I am at high genetic risk?”, and 3) the polygenic GRS was computed using selected genetic variants in all AMD biologic pathways associated with progression to higher severity stages from earlier non-advanced stages of AMD,^15,16,27,43^ rather than a GRS related to prevalence in cross-sectional data. In a previous report, analyses found that adding variants that were not associated with disease progression did not improve the effect of the GRS on risk of progression. ^15^ Although a high GRS and adherence to lifestyle habits affect the individual, using eyes as the unit of analyses increased the statistical power of the study and enabled the control for eye-specific covariates, most importantly baseline AMD severity and fellow eye status, which can vary between the two eyes of an individual. ^45,46^ Limitations of this study include the self-reported dietary intake at baseline, however the restriction of follow-up time to 5 years minimized the effect of dietary and other lifestyle changes. Other lifestyle behaviors such as physical activity, are related to AMD ^6,47^ but this information was not available in the AREDS cohort. Results may not be generalizable to other populations since participants in clinical trials are generally well motivated, more educated and well-nourished than the general population. ^21^

### Conclusions

Based on the results of this study, it is high priority to identify people with high genetic risk who have a much greater risk of progressing to AAMD and therefore have the highest potential gain from lifestyle modifications. Although the advice to adhere to healthy lifestyle applies to all individuals with AMD, these results underscore the importance of targeted lifestyle interventions in individuals with heightened risk, to reduce progression from early and intermediate AMD to advanced debilitating stages. Therefore, individuals with a strong family history of AMD should not assume they are pre-destined to develop AMD. In fact, it was estimated in this study that more than half of AAMD cases could be prevented among such individuals by modifying lifestyle behaviors: smoking cessation, maintaining a healthy BMI and restricting caloric consumption to include nutrient rich food groups such as green leafy vegetables and fish, foods rich in lutein/zeaxanthin and omega-3 fatty acids.

### Clinical Relevance

Currently detection of earlier stages of AMD is usually based on routine examinations or is an incidental finding in individuals with no or unrelated symptoms. Symptoms of reduced visual function may accompany the onset of intermediate forms of the disease. These stages can lead to advanced AMD with visual impairment and legal blindness in elderly individuals who may face other difficulties such as accessing and affording health care and insurance coverage, functional and physical co-morbidities and lower health related quality of life. ^48^ Since modifiable factors are important to be addressed to prevent progression even with a high genetic susceptibility, patient education regarding these important lifestyle choices is essential. The future management of early and intermediate AMD could integrate genetic and lifestyle data ^49,50^ and include cost-effective genetic screening and risk prediction tools to help identify at-risk individuals. Clinicians can reassure and educate patients and their concerned family members that proactive lifestyle changes may help mitigate progression to an advanced stage of AMD and loss of vision.

## Data Availability

All data produced are available online at National Institutes of Health Database of Genotype and Phenotypes through accession number phs000001.v3.p1

https://www-ncbi-nlm-nih-gov.umassmed.idm.oclc.org/projects/gap/cgi-bin/study.cgi?study_id=phs000001.v3.p1

---

**Supplemental Table 1.** Association of Progression to GA by Baseline AMD severity group and GRS tertile over 5 years.

|  |  |  |  |  |  |  |  |
| --- | --- | --- | --- | --- | --- | --- | --- |
|  | GRS Tertiles |  |  |  |  |  | Total eyes |
|  | Tertile 1 (Low Genetic Risk)<br>0.263-1.458 |  | Tertile 2 (Medium Genetic Risk)<br>1.459-1.881 |  | Tertile 3 (High Genetic Risk)<br>1.882-3.108 |  |  |
| Baseline AMD Severity Group | Total | Progressing eyes,<br>(Progression Rate to GA) | Total | Progressing eyes,<br>(Progression Rate to GA) | Total | Progressing eyes,<br>(Progression Rate to GA) |  |
| Group 2 (early AMD) | 633 | 3 (0%) | 463 | 2 (0%) | 309 | 3 (1%) | 1405 |
| Group 3 (intermediate AMD) | 266 | 29 (11%) | 437 | 41 (9%) | 589 | 90 (15%) | 1292 |
|  | 899 | 32 (4%) | 900 | 43 (5%) | 898 | 93 (10%) | 2697 |

AMD = age related macular degeneration; GA = geographic atrophy; GRS = genetic risk score

**Supplemental Table 2.** Association of Progression to NV by Baseline AMD severity group and GRS tertile over 5 years.

|  |  |  |  |  |  |  |  |
| --- | --- | --- | --- | --- | --- | --- | --- |
|  | GRS Tertiles |  |  |  |  |  | Total eyes |
|  | Tertile 1 (Low Genetic Risk)<br>0.263-1.458 |  | Tertile 2 (Medium Genetic Risk)<br>1.459-1.881 |  | Tertile 3 (High Genetic Risk)<br>1.882-3.108 |  |  |
| Baseline AMD Severity Group | Total | Progressing eyes,<br>(Progression Rate to NV) | Total | Progressing eyes,<br>(Progression Rate to NV) | Total | Progressing eyes,<br>(Progression Rate to NV) |  |
| Group 2 (early AMD) | 633 | 3 (0%) | 463 | 6 (1%) | 309 | 18 (6%) | 1405 |
| Group 3 (intermediate AMD) | 266 | 13 (5%) | 437 | 47 (11%) | 589 | 100 (17%) | 1292 |
|  | 899 | 16 (2%) | 900 | 53 (6%) | 898 | 118 (13%) | 2697 |

AMD = age related macular degeneration; GRS = genetic risk score; NV = neovascular

**Supplemental Table 3.** Cross-tabulation of family history of AMD and GRS tertile.

|  |  |  |  |  |  |
| --- | --- | --- | --- | --- | --- |
|  |  | GRS Tertiles |  |  | Total eyes |
|  |  | Tertile 1 (Low Genetic Risk)<br>0.263-1.458 | Tertile 2 (Medium Genetic Risk)<br>1.459-1.881 | Tertile 3 (High Genetic Risk)<br>1.882-3.108 |  |
| Family History of AMD | No family member affected, n (%) | 726 (38) | 636 (34) | 531 (28) | 1893 |
|  | 1 family member affected, n (%) | 123 (23) | 188 (34) | 236 (43) | 547 |
|  | ≥ 2 family members affected, n (%) | 50 (19) | 76 (30) | 131 (51) | 257 |
| Total eyes |  | 899 | 900 | 898 | 2697 |

AMD = age related macular degeneration; GRS = genetic risk score;
All percentages are row %
Odds Ratio for GRS tertile 3 vs. tertile 1 comparing subjects with ≥ 2 family members affected vs. none = 3.6, 95% CI 2.3-5.6, P<0.001

**Supplemental Table 4.** Association between progression to AAMD and AREDS treatment a over 5 years and risk-inducing profile among eyes with high genetic risk according to smoking status N = 152 events/684 eyes.

| Supplemental Table 4. Association between progression to AAMD and AREDS treatment <sup>a</sup> over 5 years and risk-inducing profile among eyes with high genetic risk according to smoking status |  |  |  |  |
| --- | --- | --- | --- | --- |
| N = 152 events/684 eyes |  |  |  |  |
| Lifestyle Profile | Never Smokers<br>n = 58 events/286 eyes |  | Ever Smokers<br>n = 94 events/398 eyes |  |
|  | HR (95% CI) <sup>b</sup> | P-value | HR (95% CI) <sup>b</sup> | P-value |
| Ideal health-promoting profile <sup>c</sup> | 1.0 (Ref) | - | 1.0 (Ref) | - |
| High risk-inducing profile <sup>d</sup> | 3.4 (1.6, 7.1) | 0.002 | 5.8 (2.1, 15.7) | <0.001 |

AAMD = advanced age-related macular degeneration; AREDS = Age-Related Eye Disease Study; BMI = body-mass index; HR = Hazard Ratio; CI = Confidence Interval
<sup>a</sup> Treatment group consists of subjects treated by antioxidants only, zinc only, or a combination of antioxidants and zinc.
<sup>a</sup> A health-promoting profile for never smokers consists of BMI < 25, caloric intake/day ≤ sex-specific median, green leafy vegetables ≥ 2.7 servings/week and fish intake ≥ 2 servings/week; for ever smokers the profile also includes past smoking
<sup>b</sup> A risk-inducing profile for never smokers consists of BMI ≥ 25, caloric intake/day > sex-specific median, green leafy vegetables < 2.7 servings/ week and fish intake < 2 servings/week; for ever smokers the profile also includes current smoking green leafy vegetables < 2.7 servings/ week and fish intake < 2 servings/week; for ever smokers it in addition consists of current smoking and all of the previous behaviors

**Supplemental Table 5.**
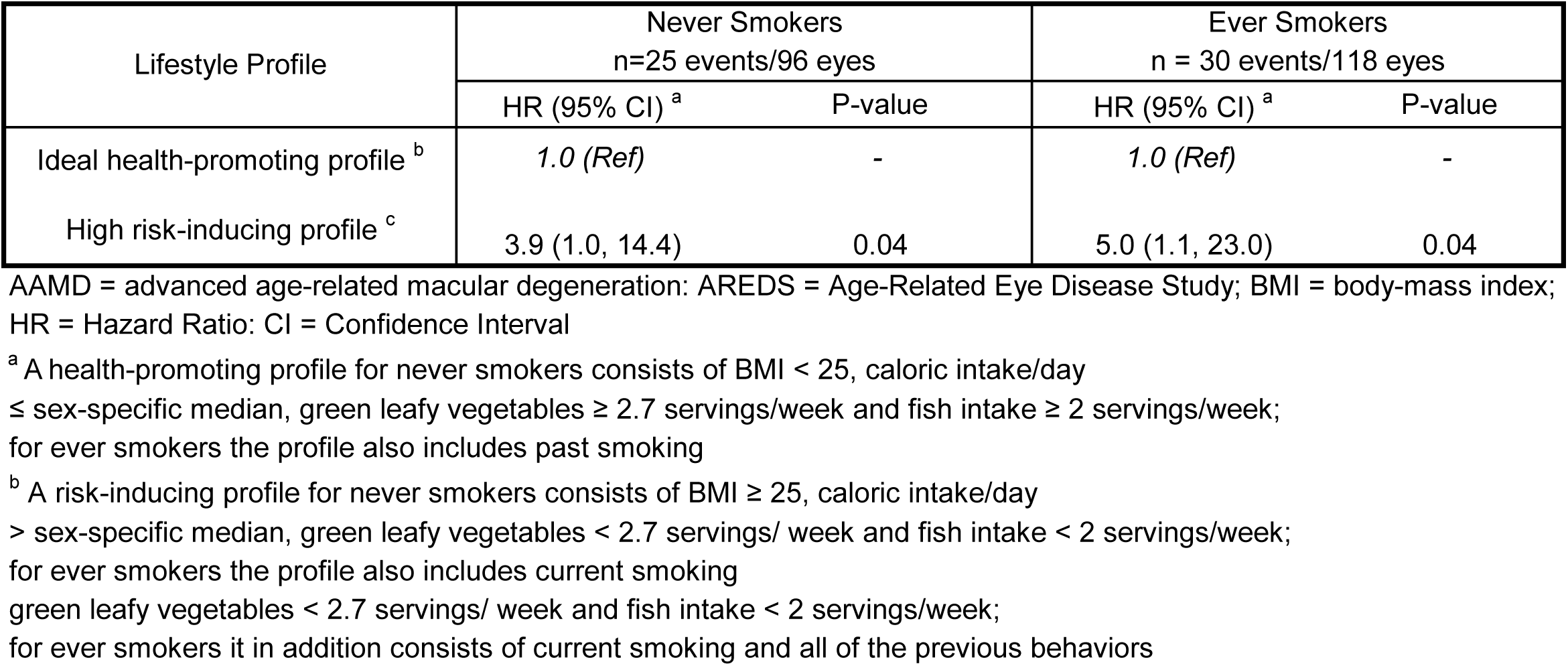
Association between progression to AAMD and AREDS placebo over 5 years and risk-inducing profile among eyes with high genetic risk according to smoking status N = 55 events/214 eyes.

**Supplemental Table 6.**
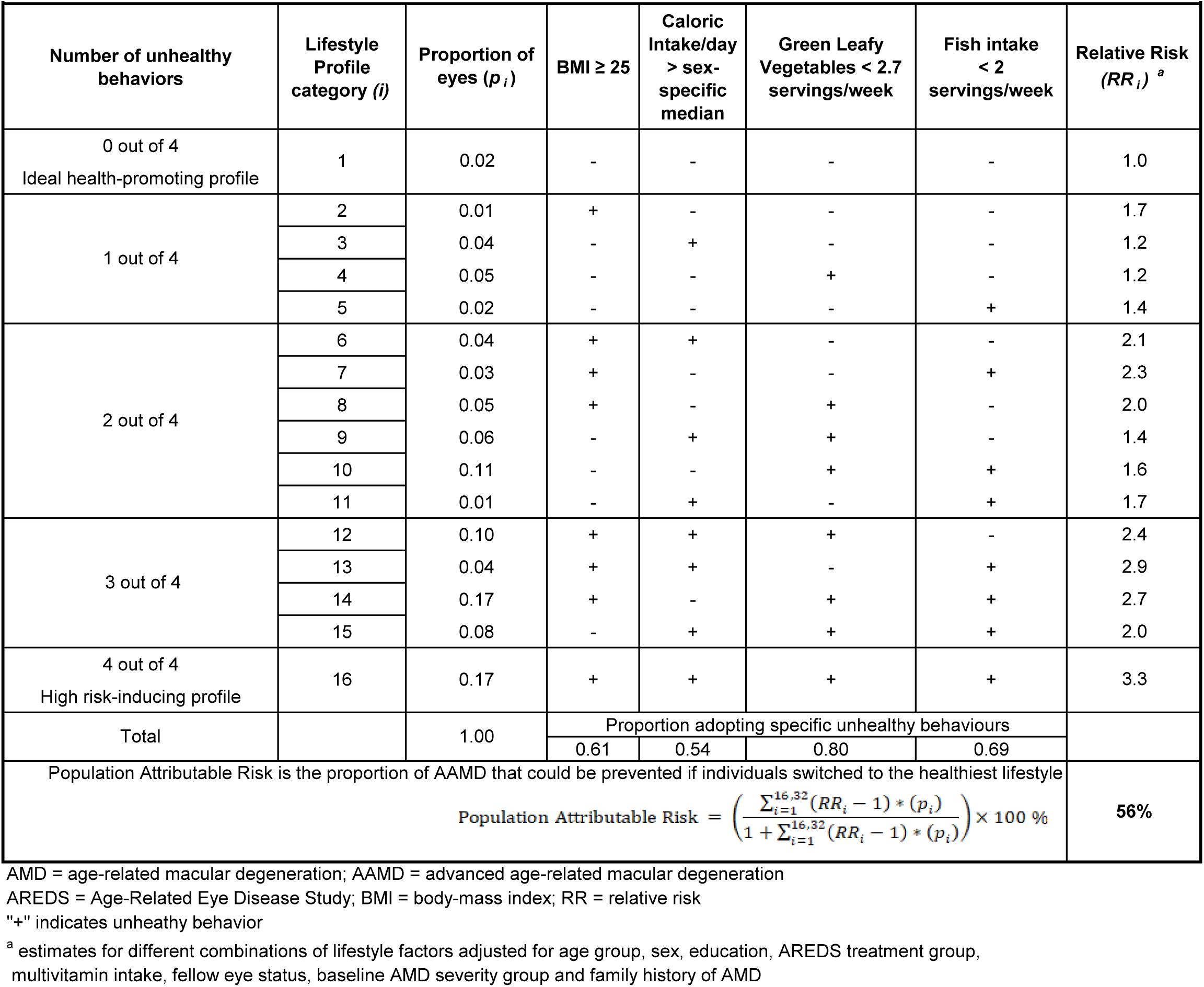
Population Attributable Risk associated with different lifestyle profile categories and progression to AAMD in never smokers with high genetic risk (N=83 events/382 eyes)

**Supplemental Table 7.**
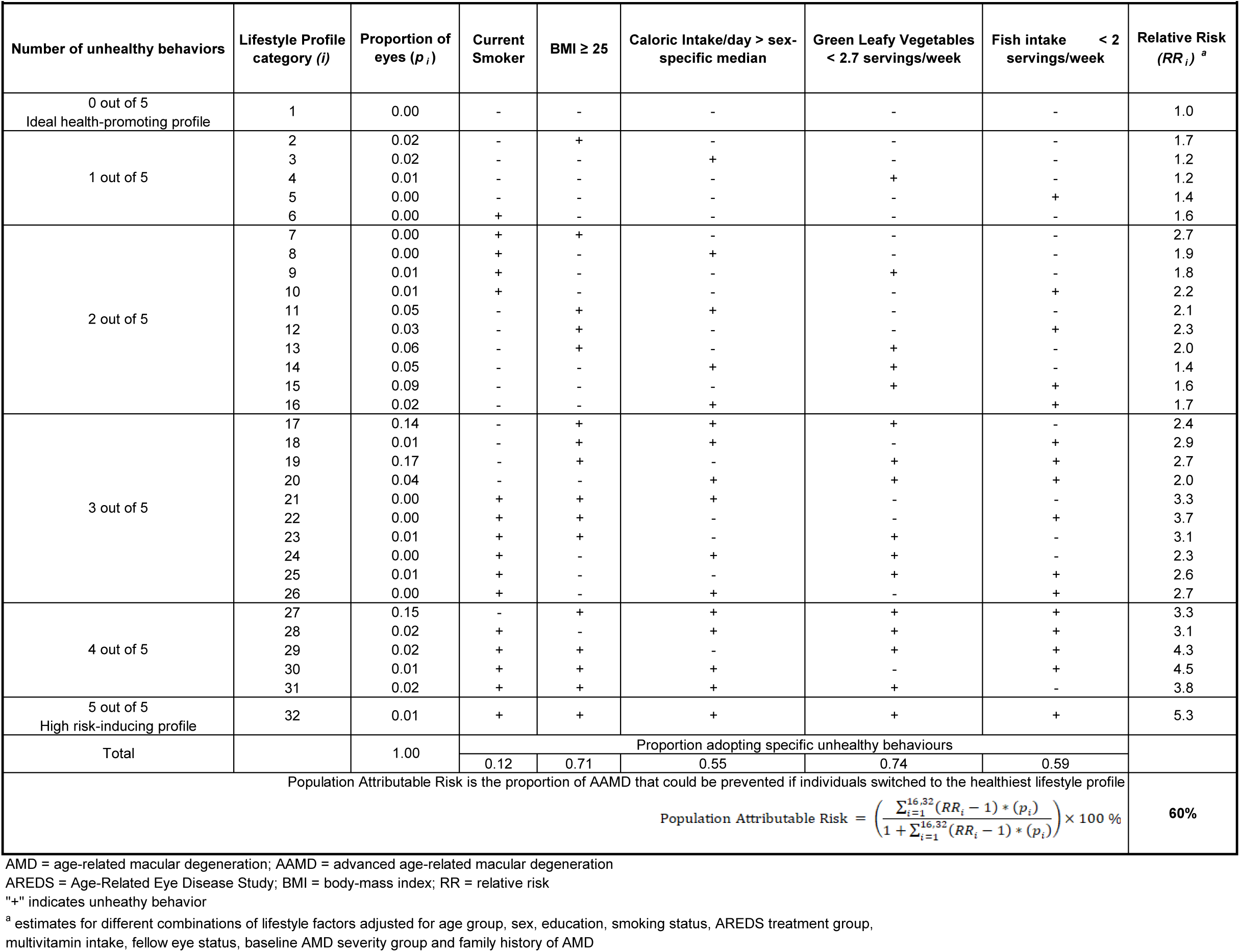
Population Attributable Risk associated with different lifestyle profile categories and progression to AAMD in ever smokers with high genetic risk (n=124 events/516 eyes)

**Supplemental Figure 1.**
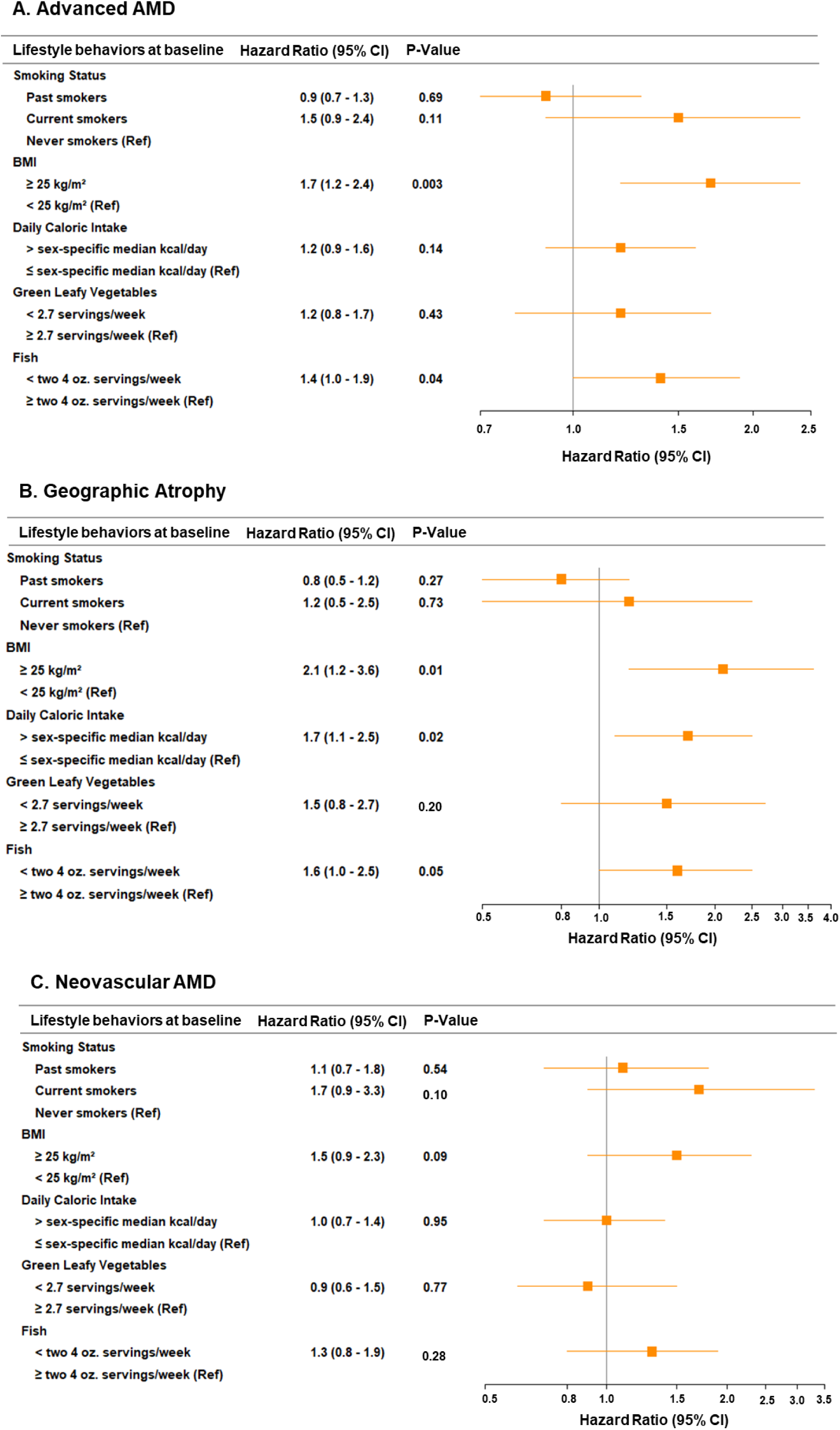
Multivariate associations between lifestyle behaviors at baseline and progression to A) Advanced AMD, B) Geographic Atrophy, and C) Neovascular AMD over 5 years in eyes with high genetic risk. Estimates adjusted for age group, sex, education, AREDS treatment group, multivitamin intake, fellow eye status, baseline AMD severity group and family history of AMD. AMD = age-related macular degeneration; CI = Confidence Interval; oz. = ounces; Ref = Reference

## SUPPLEMENTAL APPENDIX

### I. Genetic Risk Score (GRS) for progression = 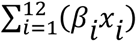

where *β*_*i*_ is the regression coefficient for the number of risk alleles present (*x*_*i*_) for 12 loci from 9 genes, adjusted for age, race and sex.

GRS = 0.06284*rs1061170_al (C) + 0.37898*rs10490924_al (T) + 0.35165*rs1410996_al (C) + 0.77838*rs121913059_al (T) + 0.11902*rs2230199_al (G) + 0.47876*rs147859257_al + 0.07387*rs8017304_al (A) + 0.15860*rs334353_al (T) + 0.10435*rs1883025_al (T) + 0.11990*rs9542236_al (C) + 0.24374*rs9621532_al (A) + 0.11180*rs8135665_al (T)

The variable name signifies the number of risk alleles per genetic variant (the risk allele is in parenthesis). The protective variants were recoded such that each component of the GRS had a positive association with progression.

### II. Lifestyle Profile for never smokers 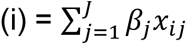

where *β_j_* is the regression coefficient for the *j*^th^ lifestyle behavioral variable (high BMI, high daily caloric intake, low weekly intake of green leafy vegetable and low weekly intake of fish) and *x_ij_* is the value of the *j*^th^ variable for the *i*^th^ subject.

Lifestyle Profile for never smokers = I (BMI ≥ 25) * 0.51960 + I (Caloric Intake > sex specific median/day) * 0.20464 + I (Intake of green leafy vegetables < 2.7 servings/week) * 0.15059 + I (Intake of fish < two 4 oz servings/week) * 0.33085

Where I (a) = 1 if true; = 0 if otherwise

### III. Lifestyle Profile for ever smokers 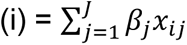

where *β_j_* is the regression coefficient for the *j*^th^ lifestyle behavioral variable (high BMI, high daily caloric intake, low weekly intake of green leafy vegetable, low weekly intake of fish and current smoking) and *x_ij_* is the value of the *j*^th^ variable for the *i*^th^ subject.

Lifestyle Profile for ever smokers = I (Current Smoking) * 0.39516 + I (Past Smoking) * (−0.06096) + I (BMI ≥ 25) * 0.51960 + I (Caloric Intake > sex specific median/day) * 0.20464 + I (intake of green leafy vegetables < 2.7 servings/week) * 0.15059 + I (Intake of fish < two 4 oz servings/week) * 0.33085

Where I (a) = 1 if true; = 0 if otherwise

### IV. Composite Risk Score = 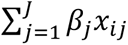

where *β_j_* is the regression coefficient for the *j*^th^ variable included in Table 2 and *x_ij_* is the value of the *j*^th^ variable for the *i*^th^ subject. This composite risk score was plotted by quartile and progression status.

Composite Risk Score = I (age65to74) * 0.62153 + I (age75plus) * 0.95458 + I (female_sex)* 0.01466 + I (education_geHS) * 0.09136 + I (areds_supplement) * (−0.13577) + I (multivitamin_yes) * (−0.02261) + I (fellow_eye_advanced) * 0.90863 + I (baselineAMD_sevgrp3)* 1.54061 + I (famhx_AMD_1) * 0.02548 + I (famhx_ge_2) * 0.25645 + I (Current Smoking) * 0.39516 + I (Past Smoking) * (−0.06096) + I (BMI ≥ 25) * 0.51960 + I (Caloric Intake > sex specific median/day) * 0.20464 + I (Intake of green leafy vegetables < 2.7 servings/week) * 0.15059 + I (Intake of fish < two 4 oz servings/week) * 0.33085

Where I (a) = 1 if true; = 0 if otherwise

